# Endogenous interferon-beta but not interferon-alpha or interferon-lambda levels in nasal mucosa predict clinical outcome in critical COVID-19 patients independent of viral load

**DOI:** 10.1101/2021.03.23.21253748

**Authors:** Soraya Maria Menezes, Marcos Braz, Veronica Llorens-Rico, Joost Wauters, Johan Van Weyenbergh

## Abstract

Although the subject of intensive preclinical and clinical research, controversy on the protective vs. deleterious effect of interferon (IFN) in COVID-19 remains. Some apparently conflicting results are likely due to the intricacy of IFN subtypes (type I: IFN-alpha/beta, type III: IFN-lambda), timing and mode of administration (nebulized/subcutaneous) and clinical groups targeted (asymptomatic/mild, moderate, severe/critical COVID-19). Within the COntAGIouS (COvid-19 Advanced Genetic and Immunologic Sampling) clinical trial, we investigated endogenous type I and type III IFNs in nasal mucosa as possible predictors of clinical outcome in critical patients, as well as their correlation to SARS-CoV-2 viral load, using nCounter technology. We found that endogenous IFN-beta expression in the nasal mucosa predicts clinical outcome, independent of viral replication or Apache II score, and should be considered as a prognostic tool in a precision medicine approach of IFN therapy in COVID-19 clinical management.

## INTRODUCTION

Although the subject of intensive preclinical and clinical research, controversy on the protective vs. deleterious effect of interferon (IFN) in COVID-19 remains. Some apparently conflicting results are likely due to the intricacy of IFN subtypes (type I: IFN-alpha/beta, type III: IFN-lambda), timing and mode of administration (nebulized/subcutaneous) and clinical groups targeted (asymptomatic/mild, moderate, severe/critical COVID-19).

Two recent phase-2 clinical trials^1,2^ reporting the use of type I and type III IFN achieved their primary clinical and virological outcomes in hospitalized and ambulatory COVID-19 patients, respectively. As set forth previously^3^, understanding the different kinetics of endogenous IFN production in mild and severe COVID-19 patients, relative to viral replication, will help identify the therapeutic window. Thus, endogenous IFN(s) add another layer of complexity to the COVID-19 IFN conundrum but have been understudied in critical (ICU) patients.

## PATIENTS AND METHODS

The COntAGIouS trial (COvid-19 Advanced Genetic and Immunologic Sampling; an in-depth characterization of the dynamic host immune response to coronavirus SARS-CoV-2) proposes a transdisciplinary approach to identify host factors resulting in hyper-susceptibility to SARS-CoV-2 infection. Within this prospective clinical trial (NCT04327570), we investigated viral and host transcriptomes in the nasal mucosa of patients with COVID-19 critical disease, admitted to the intensive care unit (ICU) at University Hospitals Leuven, Belgium. Nasal swabs were collected from 57 critical COVID-19 patients on admission in ICU and/or first bronchoscopy. RNA was extracted and mRNA levels for IFN-beta (*IFNB1* gene), IFN-alpha (*IFNA2* gene) and IFN-lambda (*IFNL2* and *IFNL3* genes) were quantified by nCounter technology (Nanostring) as previously described^4^. Viral load (total SARS-CoV-2 transcripts corresponding to Surface glycoprotein, Nucleoprotein, Envelope protein, Membrane protein, ORF1AB, ORF3A and ORF7A) was quantified as described^5^, using digital transcriptomics (nCounter, Nanostring) previously validated in a large cohort of acute respiratory infection^6^. All ICU patients received standard-of-care treatment (corticosteroids, anticoagulants, vasopressors and/or antibiotics, in addition to ventilation/ECMO) but none received IFN treatment.

## RESULTS AND DISCUSSION

As primary endpoint, we investigated length of stay in the ICU (ranging from 3 to 74 days) with type I and III IFN as predictors, using Kaplan-Meier curves. As shown in Fig. 1A-B-C, *IFNB1* transcript levels (Hazard ratio (HR) 0.30 95%CI[0.16-0.56], p=0.0001) but not *IFNA2* (HR 0.82 95%CI[0.46-1.50], p=0.53) or *IFNL2/IFNL3* transcript levels (HR 0.67 95%CI[0.35-1.27], p=0.22), nor viral load (HR 1.13 95%CI[0.64-1.98], p=0.68, data not shown) predicted the length of ICU stay. Multivariable regression confirmed *IFNB1* levels (β=0.45 [0.24-0.67], p=0.0002) and Acute Physiology and Chronic Health Evaluation (Apache) II score (β=1.06 [0.49-1.65], p=0.0009) as independent predictors, whereas viral load, age, gender, BMI or Charlson Comorbidity index were not. Moreover, *IFNB1* levels also predicted worse clinical outcome measured by maximal WHO ordinal scale or maximal oxygen support (Mann-Whitney, p=0.027 and p=0.0068, respectively), as well as a composite score (discharge to rehabilitation centre, hospital stay >60 days or death; Mann-Whitney p=0.040). Noteworthy, 45% (5 of 11) of *IFNB1*-positive patients required ECMO vs. only 9% (4 of 46) of *IFNB1*-negative patients. The total days on ECMO was also higher in *IFNB1*+ patients (median 24.0 vs. 10.5 days, Mann-Whitney p=0.016). *IFNB1* levels also predicted multi-organ involvement, another hallmark of critical COVID-19, as measured by Sequential Organ Failure Assessment (SOFA) score (median SOFA score 7 for *IFNB1*-negative vs. 12 for *IFNB1*+, Mann-Whitney p=0.0072). Surprisingly, *IFNB1* levels were not correlated to viral load (Fig. 1D), in contrast to *IFNA2* (r=0.45, p=0.0007) and *IFNL2/IFNL3* (r=0.47, p=0.0003).

**Fig. 1:**
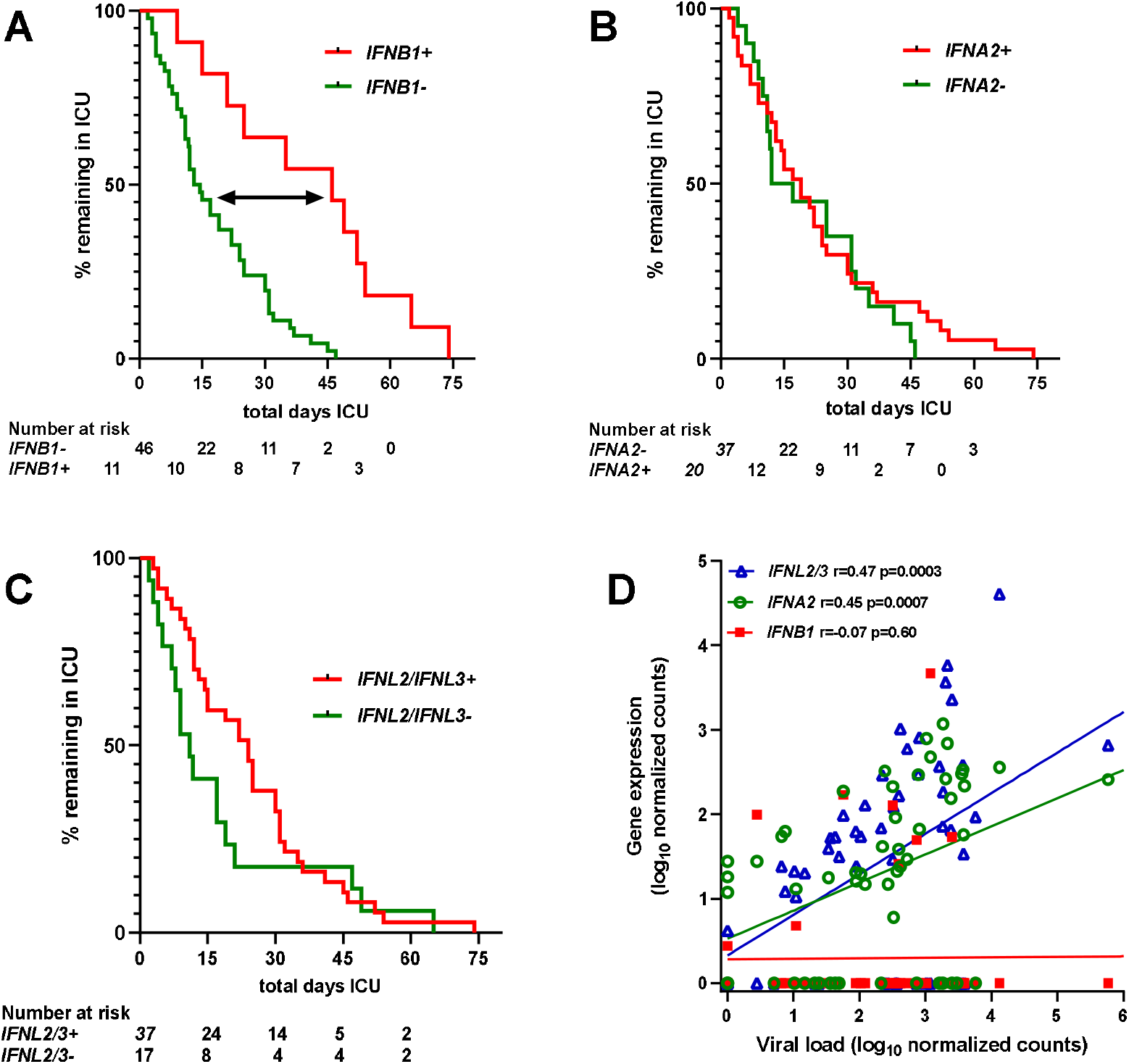
Upper respiratory tract IFN-beta transcript levels, but not IFN-alpha or IFN-lambda transcript levels, predict length of ICU stay in critical COVID-19 patients. Kaplan-Meier curves of **(A)** *IFNB1*-positive vs. *IFNB1*-negative, **(B)** *IFNA2-*positive vs. *IFNA2*-negative, and (C) *IFNL2/3-*positive vs. *IFNL2/3*-negative ICU patients were compared using Log-rank test (***p=0.0001 for *IFNB1*, not significant for *IFNA2, IFNL2/IFNL3* or viral load, not shown). (D) Viral load was correlated to *IFNB1, IFNA2*, and *IFNL2/IFNL3* transcripts (Spearman correlation). Viral load and IFNL2/IFNL3 data were missing from 3 patients (n=54).

In conclusion, endogenous IFN-beta production in the nasal mucosa predicts clinical outcome, independent of viral replication or Apache II score, and should be considered as a prognostic tool in a precision medicine approach of IFN therapy in COVID-19 clinical management.

## Data Availability

All data are available in the manuscript or upon request from the authors.

